# Investigating the causal effect of potential therapeutic agents for colorectal cancer prevention: a Mendelian randomization analysis

**DOI:** 10.1101/2025.07.07.25330867

**Authors:** Ella Fryer, D Timothy Bishop, Peter T Campbell, Andrew T Chan, Loic Le Marchand, Christopher I Li, Victor Moreno, Marc J Gunter, Amanda I Phipps, Robert C Grant, Stephanie L Schmit, Richard M Martin, James Yarmolinsky, Philip C Haycock

**Affiliations:** MRC Integrative Epidemiology Unit, University of Bristol, Bristol, England, BS8 2BN, UK; Population Health Sciences, Bristol Medical School, University of Bristol, Bristol, England, BS8 2BN, UK; Leeds Institute of Cancer and Pathology, University of Leeds, Leeds, UK; Department of Epidemiology and Population Health, Albert Einstein College of Medicine, Bronx, NY, USA; Division of Gastroenterology, Massachusetts General Hospital and Harvard Medical School, Boston, Massachusetts, USA; Channing Division of Network Medicine, Brigham and Women’s Hospital and Harvard Medical School, Boston, Massachusetts, USA; Clinical and Translational Epidemiology Unit, Massachusetts General Hospital and Harvard Medical School, Boston, Massachusetts, USA; Broad Institute of Harvard and MIT, Cambridge, Massachusetts, USA; University of Hawaii Cancer Center, Honolulu, Hawaii, USA; Public Health Sciences Division, Fred Hutchinson Cancer Center, Seattle, Washington, USA; Unit of Biomarkers and Suceptibility (UBS), Oncology Data Analytics Program (ODAP), Catalan Institute of Oncology (ICO), L’Hospitalet del Llobregat, 08908 Barcelona, Spain; ONCOBELL Program, Bellvitge Biomedical Research Institute (IDIBELL), L’Hospitalet de Llobregat, 08908 Barcelona, Spain; Consortium for Biomedical Research in Epidemiology and Public Health (CIBERESP), 28029 Madrid, Spain; Department of Clinical Sciences, Faculty of Medicine and health Sciences and Universitat de Barcelona Institute of Complex Systems (UBICS), University of Barcelona (UB), L’Hospitalet de Llobregat, 08908 Barcelona, Spain; Department of Epidemiology and Biostatistics, School of Public Health, Imperial College London, London, England, W2 1PG, UK; Department of Epidemiology, University of Washington, Seattle, Washington, USA; Division of Medical Oncology and Hematology, Princess Margaret Cancer Centre, University Health Network, Toronto, Canada; Genomic Medicine Institute, Cleveland Clinic, Cleveland, OH, USA; Department of Molecular Medicine, Cleveland Clinic Lerner College of Medicine of Case Western Reserve University School of Medicine, Cleveland, OH, USA; NIHR Bristol Biomedical Research Centre, University Hospitals Bristol and Weston NHS Foundation Trust, University of Bristol, Bristol, England, BS8 2BN, UK

## Abstract

**Background:** Conventional observational studies have identified several potential therapeutic agents that may lower risk of colorectal cancer development. However, these studies are susceptible to unmeasured and residual confounding and reverse causation, undermining robust causal inference.

**Methods:** We used Mendelian randomization (MR), a genetic epidemiological method that can strengthen causal inference, to evaluate the effect of previously reported therapeutic agents on colorectal cancer risk, including medications, dietary micronutrients, and endogenous hormones. Genetic instruments were constructed using genome-wide association studies (GWASs) of molecular traits (e.g. circulating levels of protein drug targets, blood-based biomarkers of micronutrients). Using summary statistics from these GWASs and a colorectal cancer risk GWAS (cases=78,473, controls=107,143), we employed Wald ratios and inverse-variance weighted models to estimate causal effects.

**Results:** We found evidence for associations between genetically-proxied elevated omega-3 fatty acids (OR 1.10; 95% CI 1.03, 1.18; *p*=6.20×10^-3^) and reduced plasma ACE levels (OR 1.08; 95% CI 1.03, 1.13; *p*=9.36×10^-4^), and colorectal cancer risk. Findings for ACE inhibition were consistent across sensitivity analyses and anatomical subsites.

**Conclusions:** Reduced plasma ACE levels were robustly linked to increased colorectal cancer risk. Further work is required to better understand the mechanism behind this finding and whether this translates to adverse effects via medication use (i.e. ACE inhibitors).

**Impact:** These findings provide updated evidence on the role of previously reported therapeutic agents in colorectal cancer risk, helping to prioritise further evaluation of those agents with potential aetiological roles in cancer development.

## INTRODUCTION

Colorectal cancer presents an increasing public health burden, with over 1.9 million new cases globally in 2022, predicted to rise to 3.2 million annually by 2040 (1, 2). Incidence has been increasing in low-and middle-income countries in particular, correlated with economic growth and adoption of a western lifestyle, and amongst a younger demographic (i.e. under the age of 50) in high-income countries (3–6). An individual’s risk of developing colorectal cancer is influenced by several factors, including hereditary genetic mutations and family history, which account for 5-10% of colorectal cancer cases, and a number of environmental and lifestyle factors (7–9). Whilst many of these risk factors are modifiable, such as elevated body mass index (BMI), physical inactivity, and alcohol consumption; lifestyle and behavioural changes can be difficult to implement. Screening and removal of polyps during colonoscopy in high-income countries has been an effective strategy for reducing colorectal cancer incidence and mortality in average-risk groups; however, given incomplete uptake (10–12), screening alone is insufficient to tackle overall disease burden, and additional prevention strategies are needed (13).

Therapeutic prevention is one strategy that could be used to reduce an individual’s risk of developing colorectal cancer. Therapeutic prevention involves administering a synthetic, natural, or biological agent that can prevent, reverse or delay the onset of disease (14, 15). These agents could be used for primary prevention in a healthy population (i.e. showing no signs of disease) to reduce their overall risk, or in individuals who have had polyps (a precursor to colorectal cancer) removed during colonoscopy screening, known as secondary prevention (15, 16). Examples of successful preventive therapy agents for colorectal cancer include aspirin, for which a daily dose has been shown to reduce cancer incidence (17). Given the potential adverse effects of aspirin, including gastrointestinal bleeding, aspirin is currently only recommended for groups at high-risk of developing colorectal cancer (e.g. individuals with Lynch syndrome), and so effective therapeutic agents with more favourable safety profiles for cancer prevention are needed (18). There has been great success of preventive therapy in other disease areas including cardiovascular disease; however, there are fewer examples of successful preventive therapy for cancer, in part due to the longer timeframe of cancer development and limited availability of short-term biomarkers of drug target efficacy (14, 15). For clinical trials to be conducted, significant epidemiological evidence is required to select preventive agents for testing (15). There have been many preventive agents linked to reduced risk of colorectal cancer in the observational epidemiological literature that are pharmacologically actionable (i.e. given as a supplement or drug), including dietary micronutrients, medications and exogenous hormones. However, findings from conventional observational studies can be susceptible to biases such as confounding, due to either unmeasured or imprecisely measured confounders, and reverse causation, where associations are driven by the outcome influencing the presumed exposure. It can therefore be difficult to distinguish true causal effects from spurious correlations, and therefore the suitability of these agents as intervention targets is unclear.

Mendelian randomization uses germline genetic variants to instrument exposures of interest, here dietary micronutrients, medications and hormones. As germline genetic variants are randomly allocated at meiosis and fixed from conception, conventional sources of bias such as confounding and reverse causation should be minimised, strengthening causal inference (19–21). We used Mendelian randomization to reassess these previously identified observational relationships and provide evidence to support their causal nature (19–21).

## METHODS

We used a two-sample MR framework to evaluate the causal relevance of preventive agents that have been reported to be associated with reduced colorectal cancer risk in observational studies. We have previously published a protocol detailing our methods and proposed analyses for this study (Figure 1) (22). In brief, we first conducted a literature search of reviews of therapeutic prevention and colorectal cancer risk to identify potential preventive agents. The search strategy is available in our protocol (22) and articles were included if they reviewed observational studies conducted in humans. All reported preventive agents were extracted from each review. For each agent that we identified, we attempted to generate a genetic instrument for a corresponding molecular trait (e.g. for drugs, this was circulating levels of protein drug targets; for dietary micronutrients, this was blood-based biomarkers of micronutrients; and for exogenous hormones, this was circulating levels of endogenous hormones). For instrument selection, we searched for genome-wide association studies (GWASs) using the GWAS catalogue (23), the IEU Open GWAS (24), PubMed and the preprint servers medRxiv/bioRxiv, to identify the largest GWAS conducted in individuals of European ancestry and with complete summary data available. We then conducted a two-sample MR analysis of these potential preventive agents and colorectal cancer risk, using a GWAS meta-analysis of colorectal cancer risk available in individuals of European ancestry (cases=78,473, controls=107,143). Between study heterogeneity was calculated using the *I^2^* statistic and variants with *I^2^* >65% were excluded. Effect estimates for genetically proxied micronutrients and hormones were scaled to reflect increasing levels, whilst for the protein drug targets, estimates were scaled to reflect decreasing levels, to mimic the hypothetical intervention (i.e. micronutrient supplementation, hormone replacement therapy, or inhibition of proteins by drugs). For all molecular traits found to have evidence for an association with colorectal cancer risk (defined as p<0.05), and for which we had >10 single-nucleotide polymorphisms (SNPs) in the instrument, we conducted ‘pleiotropy-robust’ methods to examine sensitivity of results to horizontal pleiotropy bias (i.e. when a genetic variant influences the outcome, either directly or indirectly, independently of the exposure). For any protein drug targets found to have an effect on colorectal cancer risk, we performed genetic colocalisation analyses using the ‘coloc’ package in R (25), to determine if there is a shared causal variant, which is necessary, although not sufficient, to infer a causal relationship between these traits,. The posterior probabilities for a number of configurations were calculated: H0 = neither the protein drug target or colorectal cancer has a genetic association in the region, H1 = only the protein drug target has a genetic association in the region, H2 = only colorectal cancer has a genetic association in the region, H3 = both the protein drug target and colorectal cancer are associated, but with different causal variants, H4 = both the protein drug target and colorectal cancer are associated and share a single causal variant. A posterior probability of ≥0.5 was used to indicate support for a configuration.

**Figure 1:**
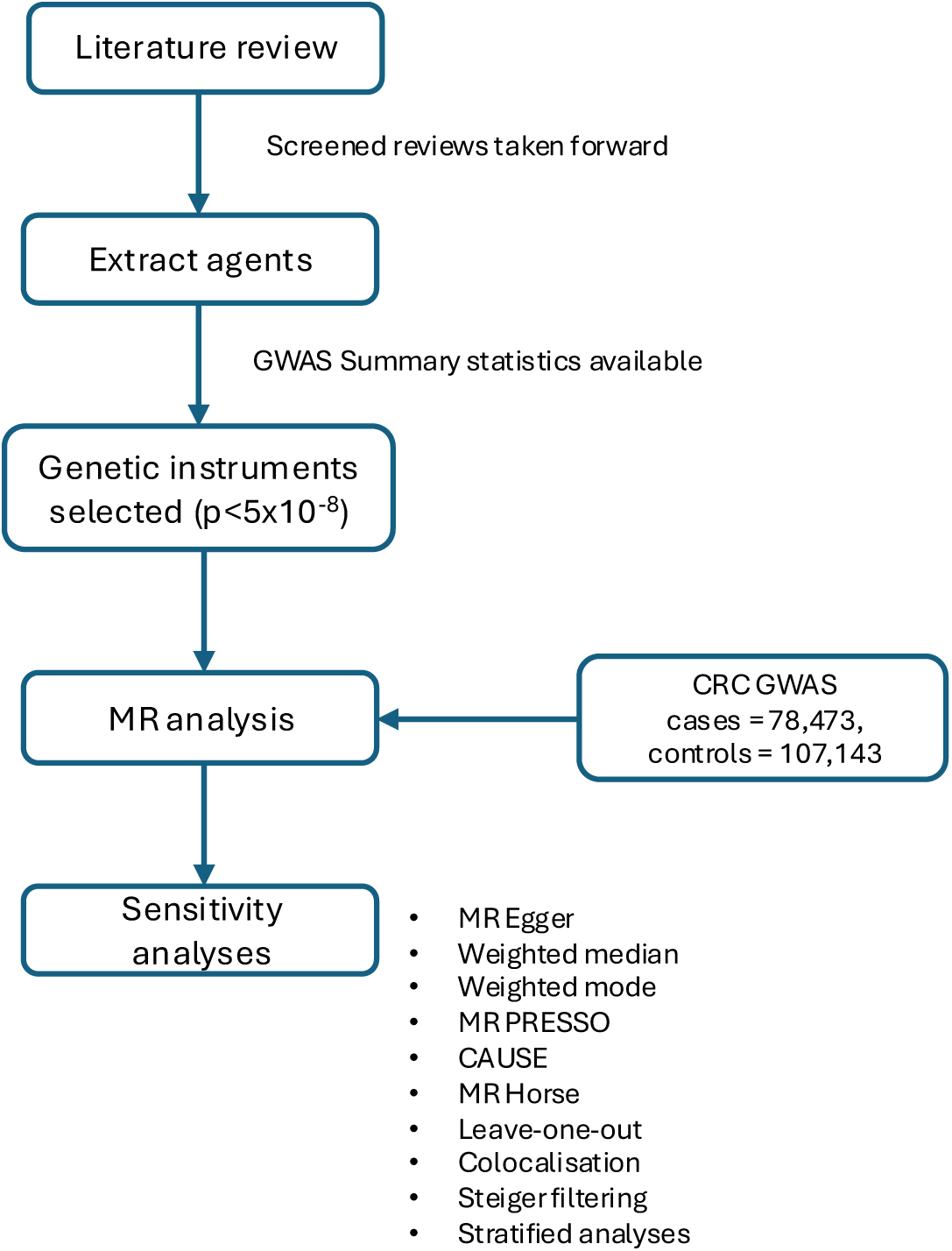
Outline of methods used in these analyses. Abbreviations: GWAS = genome-wide association study; MR = Mendelian randomization; colorectal cancer = colorectal cancer; FDR = false discovery rate; PRESSO = Pleiotropy RESidual Sum and Outlier; CAUSE = Causal Analysis Using Summary Effect estimates; Horse after the horseshoe prior.

We detail here where we have deviated from, or made additions to, our published protocol. Firstly, our protocol specified that to instrument protein drug targets and biomarkers we would allow weakly correlated SNPs (r^2^<0.1) in our genetic instrument and account for this by using a generalised IVW method (26), using a correlation matrix generated from a reference panel of 10,000 individuals from UK Biobank. To add to this, we used a ±250kb region around the cognate gene for the protein drug target to generate these instruments. We deviated from this approach when constructing instruments for ACE inhibition, where we used a region ±1MB either side of the cognate gene for the protein and an LD (linkage disequilibrium) r^2^<0.001 to select independent SNPs. This approach is consistent with previously published work, investigating the effect of genetically proxied ACE inhibition on cancer risk (27). For all other exposures (e.g. micronutrients and hormones), we used a genome-wide significance (p<5×10^-8^) threshold and an r^2^<0.001 to select instruments, as stated in the protocol. Secondly, to account for potential sex-specific genetic influences on hormone levels, we conducted MR analyses of hormone levels using colorectal cancer data stratified by sex (female: cases=26,843, controls=32,820; male: cases=31,288, controls=34,527), which was not previously stated in the protocol. For oestradiol we could only conduct analyses in men, as a genetic instrument was only available in men. Thirdly, our protocol specified that a false-discovery rate (FDR) correction of 5% was used to define strong evidence. Given that our hypotheses were guided by observational evidence, we deviated from the protocol and interpreted a p-value threshold of <0.05 as indicative of some evidence of an association, carrying forward all findings reaching this threshold for sensitivity analyses. The FDR corrected finding, along with the results of sensitivity analyses, were used to inform our interpretation of findings in the discussion. We also conducted additional sensitivity analyses for correlated horizontal pleiotropy that was not previously specified in the protocol, including MR-Horse (28) and CAUSE (29). Correlated pleiotropy occurs when the genetic variants used to instrument an exposure, affect a confounder that acts on both the exposure and outcome. CAUSE attempts to determine if genetic associations for two traits are consistent with a causal effect by comparing whether a sharing model, allowing for horizontal pleiotropic effects, or a causal model fit the data better (29). Regarding the colocalisation analyses, we used the default parameters (e.g. the prior probabilities of the SNP being associated with the exposure, the outcome or both traits is specified as 1×10^-4^, 1×10^-4^ and 1×10^-5^, respectively), as specified in the protocol. We did not specify in the protocol that ±100kb either side of the lead SNP was used to define the region and a more stringent threshold of 5×10^-6^ was used to test p12 (the SNP being associated with both traits). For any findings with evidence of an effect on colorectal cancer risk in the main analysis, we conducted MR analyses using colorectal cancer data stratified by anatomical subsite, including colon (cases=32,002, controls=64,159), rectal (cases=16,212, controls=64,159), proximal (cases=15,706, controls=64,159), and distal (cases=14,376, controls=64,159), as specified in the protocol. Additionally, we conducted analyses restricted to early age at onset (<50 years at diagnosis) (cases=6,176, controls=65,829). In addition, a z-test was used to assess differences in findings between anatomical subsites (i.e. colon and rectal, distal and proximal) and age at onset (i.e. early onset and overall), as well as differences between sexes in the hormone analysis. When performing z-tests we accounted for potential sample overlap in control groups by calculating decoupled standard errors for effect estimates (30–32).

The Strengthening the Reporting of Observational Studies in Epidemiology using Mendelian Randomization (STROBE-MR) (33) guidelines were used to structure the reporting of this study.

## RESULTS

### Literature review

We identified over 40 potential preventive agents in our literature review (Supplementary table 1). Of these, 18 had a corresponding molecular trait (e.g. levels of protein drug targets, circulating dietary biomarkers and circulating levels of endogenous hormones) that could be instrumented and for which GWAS summary statistics of direct measures of these traits were available in individuals of European ancestry (Table 1). Characteristics of genetic variants used to instrument each trait and estimates of r^2^ and F-statistic are presented in Supplementary Table 2.

**Table 1:** Potential preventive agents identified from reviews of observational studies of colorectal cancer risk and the corresponding molecular trait instrumented in Mendelian randomization analyses.

| Preventive agent | Molecular trait | GWAS PMID | Sample size |
| --- | --- | --- | --- |
| Dietary long-chain omega-3 polyunsaturated fatty acids | Circulating long-chain omega-3 polyunsaturated fatty acids | 35692035 (34) | 114999 |
| Dietary long-chain omega-6 polyunsaturated fatty acids | Circulating long-chain omega-6 polyunsaturated fatty acids | 35692035 (34) | 114999 |
| Dietary calcium | Serum calcium levels | 33462484 (35) | 313387 |
| Dietary vitamin D | Serum 25 hydroxyvitamin D | 32059762 (36) | 443734 |
| Dietary folate levels | Serum folate levels | 30339177 (37) | 2232 |
| Dietary selenium | Circulating selenium levels | 23720494 (38) | 5477 |
| Dietary vitamin A | Serum retinol levels | 38374065 (39) | 22274 |
| Dietary vitamin C | Plasma vitamin C levels | 33203707 (40) | 52018 |
| Dietary vitamin E | Circulating alpha-tocopherol levels | 36635386 (41) | 8192 |
| Dietary $\beta$ carotene | Circulating $\beta$ carotene levels | 19185284 (42) | 1190 |
| Dietary magnesium | Serum magnesium levels | 20700443 (43) | 15366 |
| Statins | LDL cholesterol levels (due to HMGCR inhibition) | 24097068 (44) | 1320016 |
|  | LDL cholesterol levels (due to NPC1L1 inhibition) | 24097068 (44) | 1320016 |
|  | LDL cholesterol levels (due to PCSK9 inhibition) | 24097068 (44) | 1320016 |
| Guselkumab | Plasma IL23 protein levels (i.e. IL23R inhibition, IL12B inhibition) | 29875488 (45) | 3301 |
| Antihypertensives | Plasma ACE levels | 37794186 (46) | 54219 |
|  | SBP (due to NCC inhibition) | 30224653 (47) | 757601 |
|  | SBP (due to ADRB1 inhibition) | 30224653 (47) | 757601 |
| Testosterone | Bioavailable testosterone levels | 32042192 (48) | 188507 (women)<br>178782 (men) |
|  | Total testosterone levels | 32042192 (48) | 230454 (women)<br>194453 (men) |
| Oestradiol | Serum oestradiol levels | 32042192 (48) | 206927 (men) |
| Sex hormone binding globulin | Serum SHBG levels | 32042192 (48) | 189473 (women)<br>180726 (men) |
| Progesterone/17-hydroxyprogesterone | Circulating progesterone levels | 34822396 (49) | 1877 (women)<br>2220 (men) |
|  | 17-hydroxyprogesterone levels | 34822396 (49) | 1329 (women)<br>2220 (men) |

*Abbreviations: GWAS = genome-wide association study; PMID = PubMed identifier; LDL = low-density lipoprotein; HMG-CoA = 3-hydroxy-3-methylglutaryl coenzyme A; NPC1L1 = NPC1 like intracellular cholesterol transporter 1; PCSK9 = proprotein convertase subtilisin/kexin type 9; IL23R = interleukin 23 receptor; IL12B = interleukin 12B; ACE = angiotensin-converting enzyme; SBP = systolic blood pressure; NCC = sodium chloride cotransporter; ADRB1 = β1 adrenoceptor; SHBG = sex hormone binding globulin. Preventive agents identified in the literature that had a corresponding molecular trait that was deemed instrumentable. These agents included dietary micronutrients, drugs and endogenous hormones. For each drug, the biological target of the drug was identified as its molecular trait, (e.g. the levels of the protein that the drug targets). For dietary micronutrients and endogenous hormones, molecular traits were identified that correspond to the agent (e.g. blood-based biomarkers for vitamin levels). For oestradiol, an instrument was only available in men.*

### Two-sample MR analyses

We took all circulating micronutrients and protein drug targets for which we could construct genetic instruments and, using MR, tested their effect on colorectal cancer risk (Figure 2) (Supplementary table 3). Effect estimates are presented as follows: odds ratios (ORs) per standard deviation (SD) unit increase/decrease in genetically proxied exposure; 95% confidence intervals; p-value.

**Figure 2:**
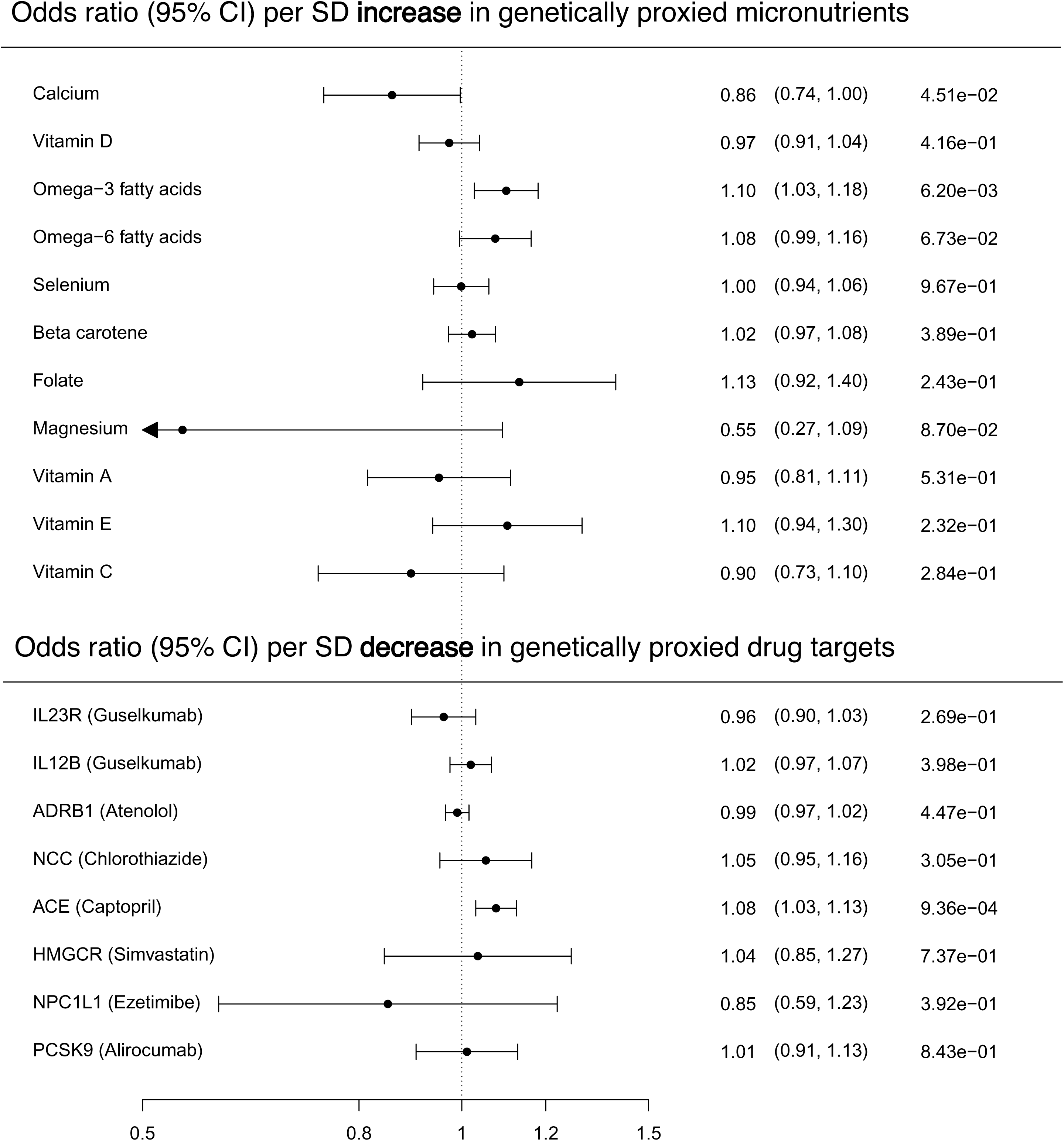
Effect estimates from initial Mendelian randomization analyses of circulating micronutrients and protein drug targets on colorectal cancer risk. Abbreviations: CI = confidence interval; SD = standard deviation; IL23R = interleukin 23 receptor; IL12B = interleukin 12B; ADRB1 = β1 adrenoceptor; NCC = ACE = angiotensin I converting enzyme; HMGCR = 3-hydroxy-3-methylglutaryl-CoA reductase; NPC1L = NPC1 like intracellular cholesterol transporter 1; PCSK9 = proprotein convertase subtilisin/kexin type 9; MR = Mendelian randomization; colorectal cancer = colorectal cancer. The odds ratio was calculated using the multiplicative random effects inverse variance weighted (IVW) method, except where the number of SNPs was limited (e.g. ≤3) in which case the fixed effects IVW was used. For those exposures instrumented by one SNP, the Wald ratio was used.

We initially found evidence that genetically proxied omega-3 fatty acids increased colorectal cancer risk (1.10; 1.03, 1.18; 6.20×10^-3^). This finding was generally consistent across ‘pleiotropy-robust’ sensitivity analyses (i.e. confidence intervals for all estimates overlapped) (Figure 3) (Supplementary Table 4) but just exceeded the FDR corrected p-value threshold (Supplementary Table 3). Leave-one-out analyses indicated that a single SNP, rs174564, may be biasing the effect estimate for omega-3 fatty acids on colorectal cancer risk, as removal of this SNP attenuated the effect to the null (1.02; 0.92, 1.13; 0.72), although confidence intervals did overlap with the overall effect estimate (Supplementary table 5). We found strong evidence that a decrease in plasma ACE levels, the mechanism of action of ACE inhibitors, increased colorectal cancer risk (OR per unit decrease in inverse-rank normalized protein expression: 1.08; 1.03, 1.13; 9.36×10^-4^) (Figure 2). There were insufficient SNPs in the genetic instrument to conduct pleiotropy-robust analyses on ACE inhibition, but this result met the FDR corrected p-value threshold. We found some evidence that circulating calcium levels had a protective effect on colorectal cancer risk (0.86; 0.74, 1.00; 0.045). Although this finding did not meet the FDR corrected p-value threshold, it was consistent across most of the pleiotropy-robust methods (Figure 3), except for CAUSE which did not find evidence for a causal relationship (Supplementary Table 6).

**Figure 3:**
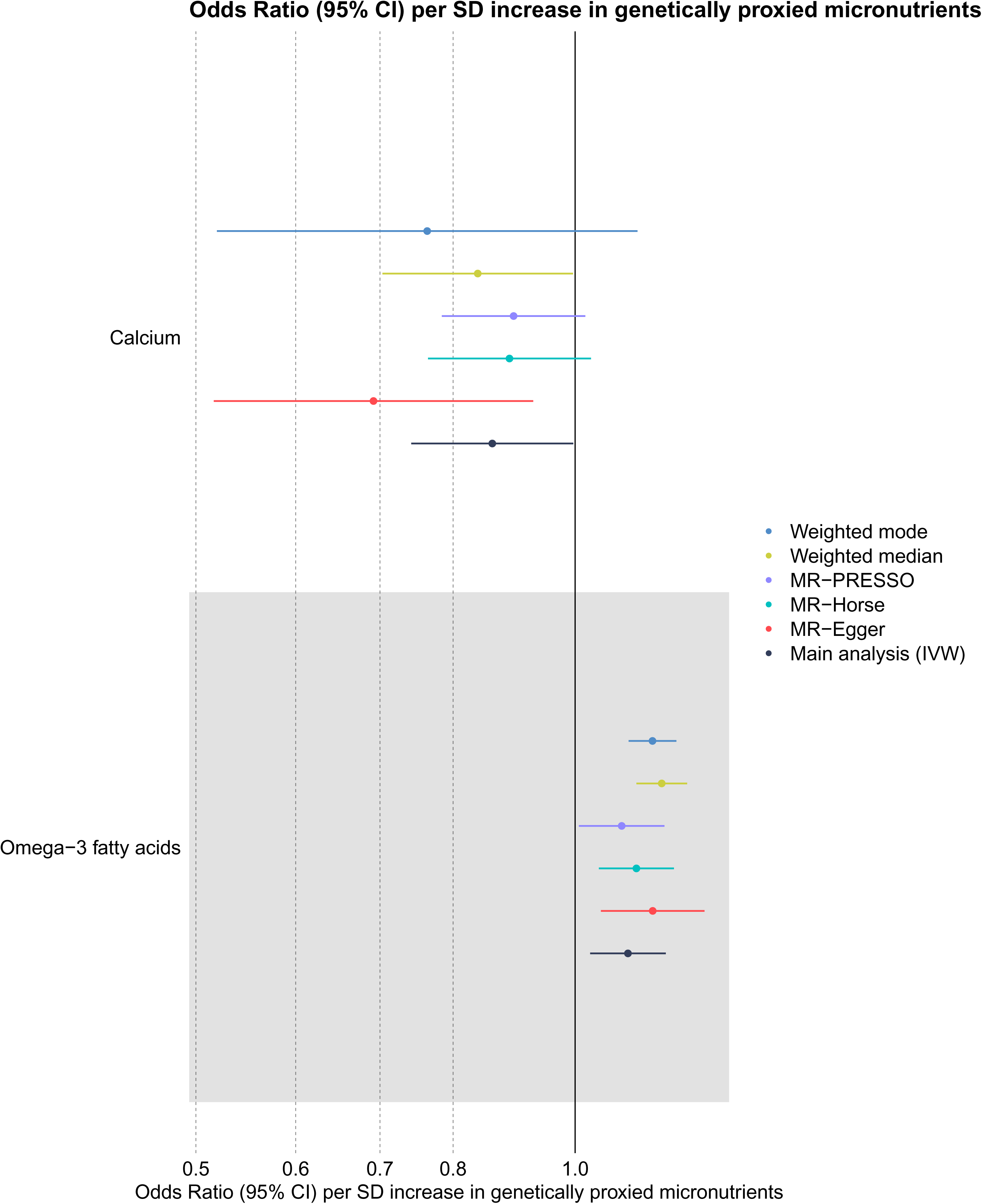
Effect estimates calculated using ‘pleiotropy robust’ methods for those micronutrients found to have an effect on colorectal cancer risk in the main analyses. Abbreviations: CI = confidence interval; SD = standard deviation; IVW = inverse variance weighted.

In analyses examining endogenous hormones we found evidence that genetically-proxied circulating progesterone had a protective effect on colorectal cancer risk in men (0.68; 0.49, 0.94; 0.019) (Figure 4) (Supplementary table 7). This finding did not meet the FDR-corrected p-value threshold and there was not strong evidence for heterogeneity between the effect estimates for men and women (z=1.719, p=0.09). We found little evidence of an effect of any other hormones on colorectal risk in either men or women. Progesterone in men was instrumented using 1 SNP, so we therefore could not conduct pleiotropy-robust sensitivity analyses.

**Figure 4:**
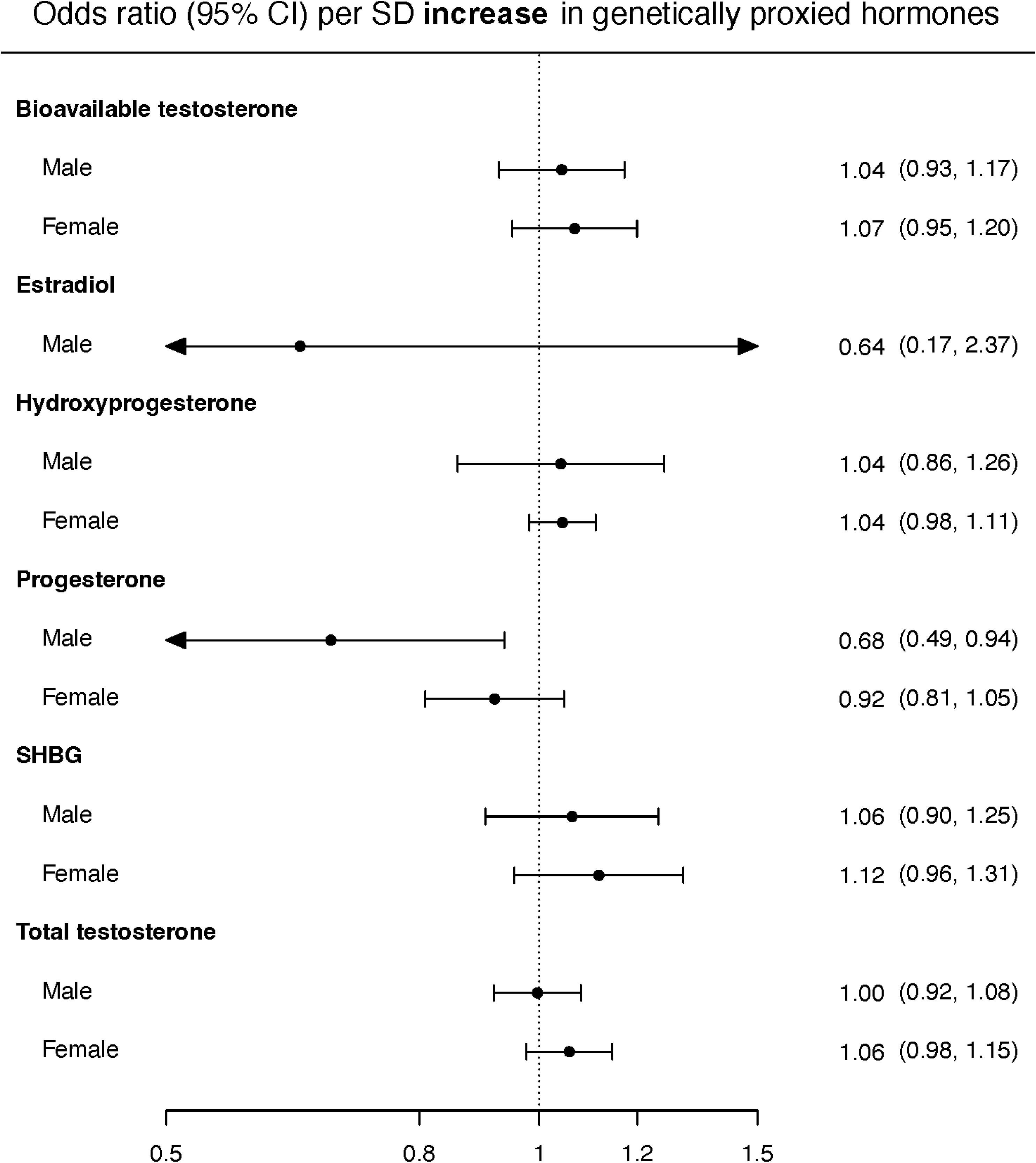
Effect estimates from Mendelian randomization analyses of circulating hormones on colorectal cancer risk in men and women. Abbreviations: SD = standard deviation; SHBG = sex hormone-binding globulin. The odds ratio was calculated using the multiplicative random effects inverse variance weighted (IVW) method, except where the number of SNPs was limited (e.g. ≤3) in which case the fixed effects IVW was used. For those exposures instrumented by one SNP, the Wald ratio was used. Effect estimates are scaled to reflect increasing levels of circulating hormones.

### Multivariable Mendelian Randomization

The univariable IVW estimated effect for omega 3 fatty acids on colorectal cancer risk was OR=1.10, 95% CIs 1.03, 1.18, p=6.20×10^-3^. Given the genetic correlation between omega-3 and omega-6 fatty acids, estimated previously using LD score regression to be over 60% (50), it can be difficult to disentangle their independent effects on an outcome. We performed multivariable MR (MVMR) analyses to estimate the effect of omega-3 fatty acids whilst adjusting for omega-6 fatty acids (Supplementary table 8). Given that the *FADS* SNP, rs174564, may have been biasing the effect estimate for omega-3 fatty acids on colorectal cancer in the main analysis, we removed this SNP from the omega-3 instrument and repeated the MVMR analysis. We also recalculated the IVW estimate for omega-3 on colorectal cancer risk with this same SNP removed (Figure 5). The MVMR effect estimate for omega-3 with rs174564 included (OR=1.07, 95% CIs=0.96, 1.18, p=0.24) and excluded (OR=1.00, 95% CIs=0.86, 1.17, p=1.00) attenuated towards the null. The univariable IVW estimate for omega-3 excluding rs174564 (OR=1.04, 95% CIs=0.95, 1.14, p=0.19) also attenuated to the null.

**Figure 5:**
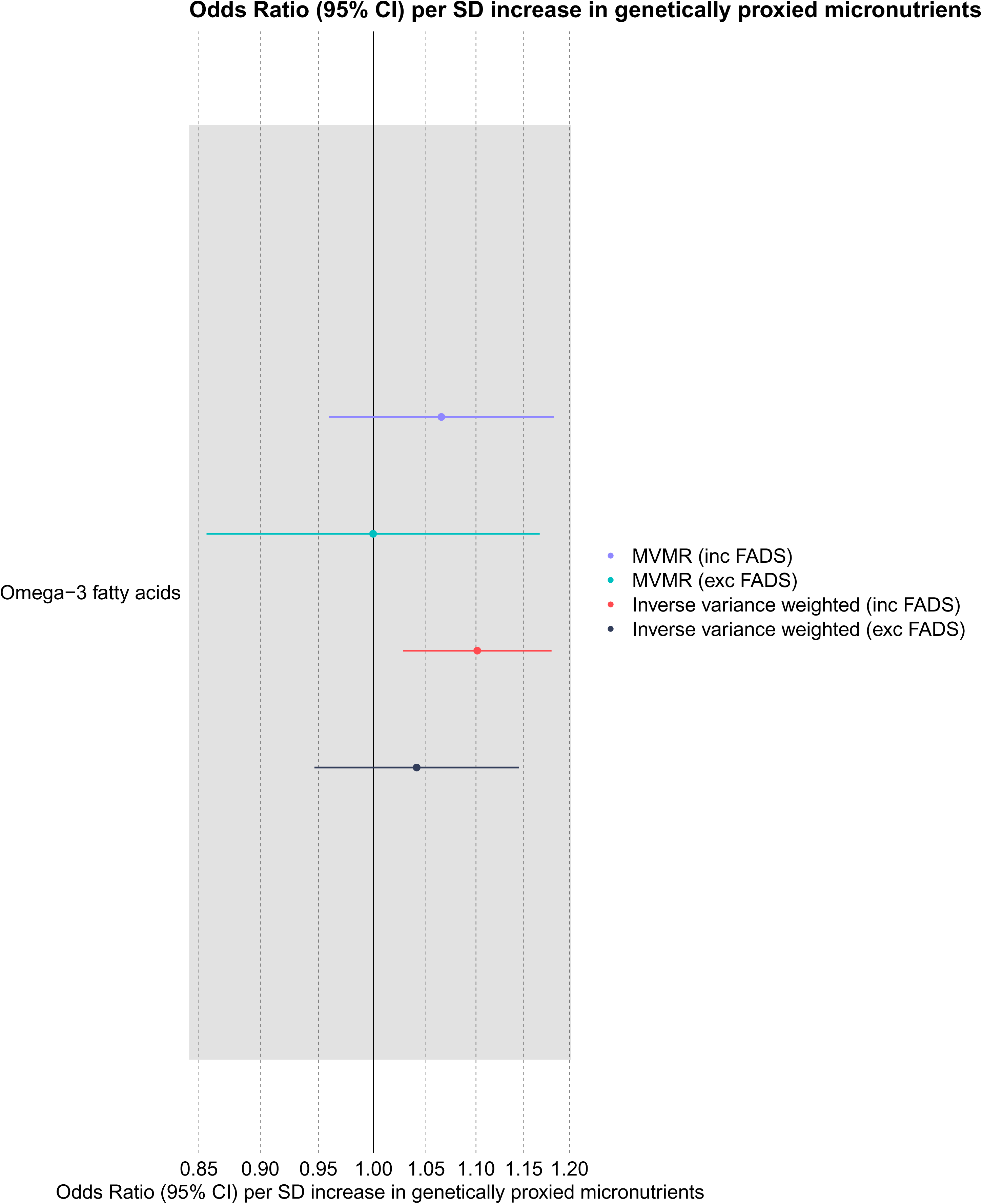
Comparison of multivariable Mendelian randomization effect estimates for fatty acids on colorectal cancer risk with the estimate from the main analyses. Abbreviations: CI = confidence interval; SD = standard deviation; MVMR = multivariable Mendelian randomization; inc = including; exc = excluding; FADS = fatty acid desaturase. This plot shows the MVMR estimates for omega-3 adjusted for omega-6 fatty acids. The omega-3 fatty acid dataset contained SNPs in the FADS gene region (e.g. rs174564), the FADS region was removed and MVMR and IVW estimates recalculated for comparison. This region was not present in the omega-6 fatty acid dataset.

### Steiger filtering

Steiger filtering can test the causal direction of SNP effects to evaluate if effect estimates are being driven by reverse causation (i.e. where the causal effect of a SNP on the exposure is mediated by the outcome). If the variance explained by a SNP was larger for the molecular trait than for colorectal cancer, this was consistent with a scenario where the molecular trait causally influences colorectal cancer risk, rather than the reverse. For both omega-3 fatty acids and calcium, all SNPs were found to explain more of the variance in the molecular trait, consistent with the molecular trait causally influencing colorectal cancer.

### Colocalisation

The colocalisation analyses of ACE inhibition and colorectal cancer risk found strong evidence that there is a shared causal variant in this region (posterior probability H4=0.94) (Figure 6) (Supplementary table 9). This finding was consistent when using a more stringent threshold for prior probability that the SNP was associated with both traits (e.g. p12=5×10^-6^) (posterior probability H4=0.89).

**Figure 6:**
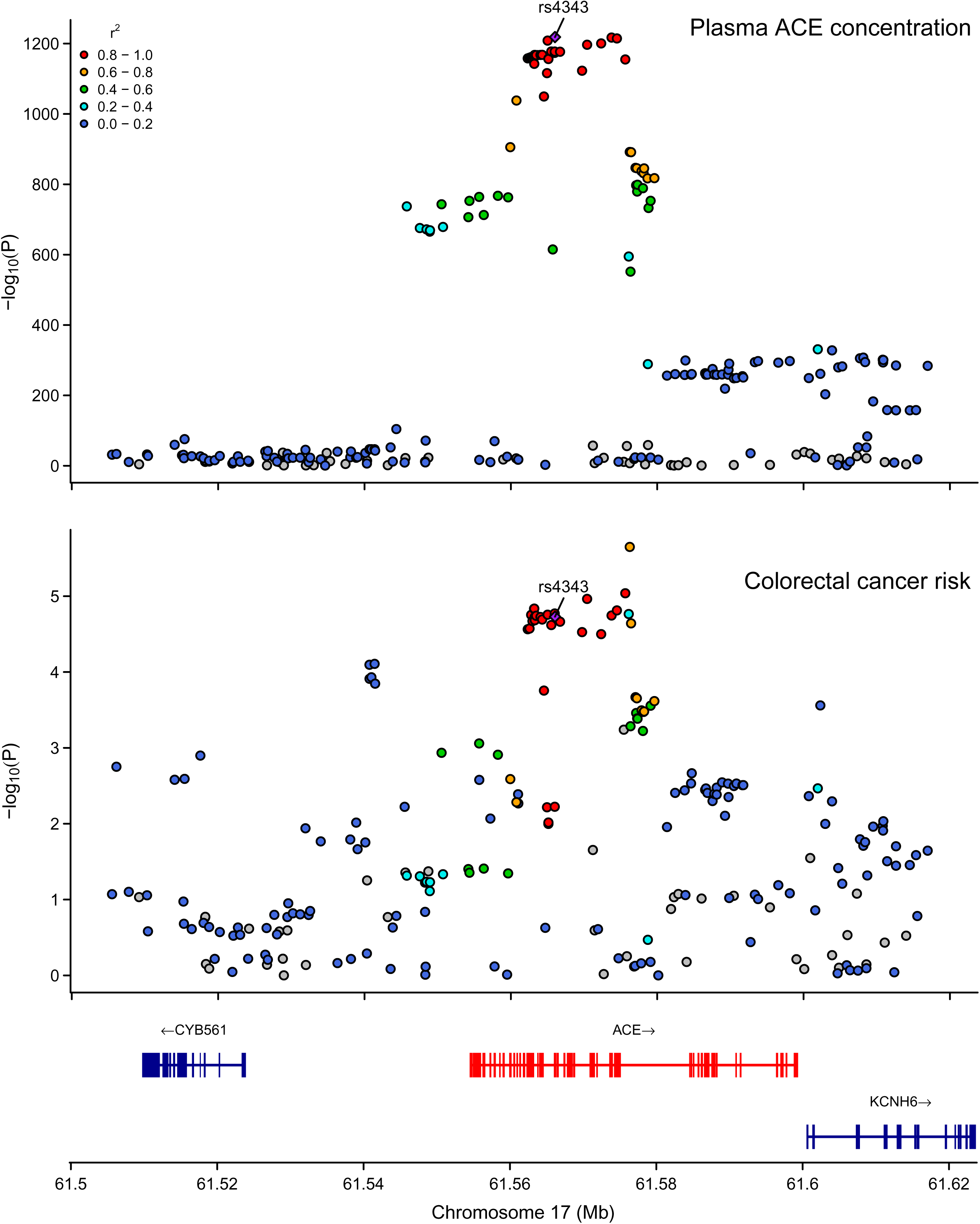
Regional association plots showing association of genetic variants with ACE inhibition and colorectal cancer risk ±100kb from the lead SNP (rs4343) used to instrument ACE. Abbreviations: ACE = angiotensin-converting enzyme; chr = chromosome; Mb = megabase; colorectal cancer = colorectal cancer. Regional association plots showing the genomic location of variants and their association (-log10(P)) with levels of ACE and colorectal cancer risk, generated using the LocusCompareR package. Posterior probability for a shared causal variant associated with both plasma ACE levels and colorectal cancer risk was 0.94, the regional association plots do not appear to support the presence of multiple independent variants driving the associations with either trait.

We attempted to perform SUSIE, which allows for multiple causal SNPs in the region, but no credible sets were found for colorectal cancer risk in the ACE locus as no SNPs in the colorectal cancer dataset reached the default minimum p-value threshold (1×10^-6^). SUSIE does not assess for evidence of an association in datasets where no SNPs fall below the minimum p-value threshold.

### Stratified analyses

For those exposures that we found evidence of an effect on overall colorectal cancer risk we also tested their effect on different anatomical subsites of colorectal cancer, including colon, rectal, proximal, and distal, and colorectal cancer that develops at an early age (i.e. <50 years old). The results were generally consistent across subsites for all molecular traits tested, with 95% CIs for all estimates overlapping (Figure 7) (Supplementary table 10).

**Figure 7:**
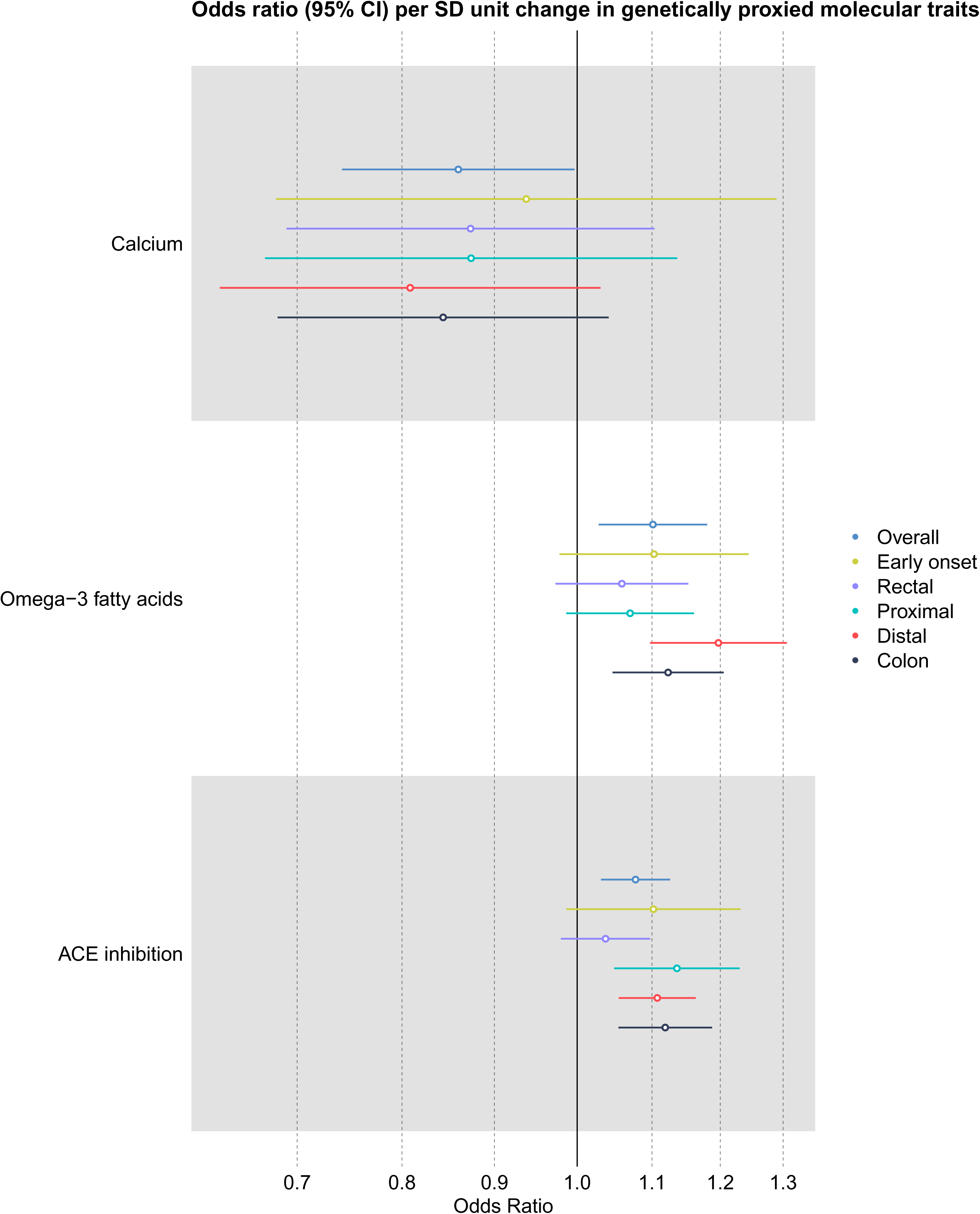
Mendelian randomization effect estimates stratified by anatomical subsite and age at onset for micronutrients and drug targets found to have an effect on overall colorectal cancer. Abbreviations: CI = confidence interval; SD = standard deviation; ACE = angiotensin converting enzyme. Effect estimates are scaled to reflect increasing micronutrient levels and decreasing protein levels.

## DISCUSSION

This Mendelian randomization analysis investigated the effects of genetically proxied circulating micronutrients, hormone levels, and inhibition of protein drug targets on colorectal cancer risk. To our knowledge, this is the largest and most comprehensive appraisal of previously reported therapeutic agents for colorectal cancer prevention, conducted in individuals of European ancestry using Mendelian randomization. We found strong evidence that genetically proxied inhibition of ACE increased colorectal cancer risk. We initially found evidence that genetically proxied elevated circulating omega-3 fatty acids increased colorectal cancer risk, however further analysis suggested other factors may have been driving this effect. We also found weak evidence for a protective effect of genetically proxied elevated circulating calcium levels.

We found strong evidence that genetically proxied ACE inhibition had an adverse effect on colorectal cancer risk, supported by genetic colocalisation of these traits around the *ACE* gene region. This finding is consistent with a previously published MR study investigating the effect of genetically proxied inhibition of antihypertensive drugs targets and a number of cancers, including colorectal cancer (27). The latter study allowed correlated SNPs in the ACE instrument (r2 <0.1) located in and within 100kb from the *ACE* gene (resulting in 14 SNPs) and used a smaller GWAS for colorectal cancer risk (cases=58,221, controls=67,694). The implications of this finding are complicated to interpret as ACE has a number of functions and effects in the body, making it difficult to pinpoint the mechanism of increasing cancer risk (51). Given that cancer is not the indication for ACE inhibitors, there have not been any RCTs of ACE inhibitor use and colorectal cancer risk specifically. However, a meta-analysis of RCTs of ACE inhibitor use (including 70 trials with 324,168 participants) that had reported overall cancer incidence as a secondary outcome, did not report evidence for an effect on cancer (fixed-effect OR: 1.00, 95% CIs: 0.92-1.09, *I^2^*=2.0%) (52). Given that this study looked at overall cancer, the findings may not be reflective of the effect of ACE inhibitor use on colorectal cancer development specifically. Our finding here provides evidence for a causal relationship between genetically proxied ACE inhibition and colorectal cancer risk, and further work is required to better understand the mechanism behind this relationship.

Genetically-proxied higher levels of circulating omega-3 fatty acids were found to have an adverse effect on colorectal cancer risk which is inconsistent with previous meta-analyses of observational studies that have found an inverse association of higher omega-3 fatty acids on colorectal cancer risk (53–56). However, it can be difficult to disentangle the effects of omega-3 and omega-6 fatty acids in MR analyses given the large genetic overlap between the fatty acids. This was demonstrated by the MVMR analyses finding attenuated effect estimates for omega-3 fatty acids when adjusting for omega-6. In addition to this, in the leave-one-out analysis, a single SNP, rs174564, was identified as driving the MR effect estimate. After removing rs174564 from the analysis, little effect of omega-3 on colorectal cancer risk was found. This SNP is located in the fatty acid desaturase (*FADS*) gene region, suggesting that activity of this enzyme accounts for the effect on colorectal cancer risk. As the desaturase enzymes encoded by the *FADS* genes are involved in rate-limiting steps in the biosynthesis of both omega-3 and omega-6 fatty acids, it is not clear if the effect of *FADS* variants on colorectal cancer risk is independent of omega-6. Previous work has looked at individual omega-3 fatty acids and colorectal cancer risk and also found little evidence for an effect after removing SNPs from the *FADS* region, including for overall increased omega-3 fatty acids (OR: 1.11, 95% CIs: 0.97-1.28, p=0.13) (32). An RCT found little evidence for an effect of supplementation with omega-3 fatty acid, eicosapentaenoic acid (EPA), on risk of having any colorectal adenomas (RR: 0.98, 95% CIs: 0.87-1.12), although secondary analyses did identify an effect on reduction of number of colorectal adenomas (incidence rate ratio (IRR): 0.86, 95% CIs: 0.74-0.99) (57).

We found weak evidence that genetically proxied circulating calcium levels had a protective effect on colorectal cancer risk and that the direction of effect remained consistent across sensitivity analyses. The modest effect we observed in this study was consistent across anatomical subsites, so it appears that this effect was not driven by an effect at a particular site. There has been some evidence from several systematic reviews and meta-analyses of randomized controlled trials (RCTs) that calcium supplementation has a modest preventive effect on recurrent colorectal adenomas. One systematic review and meta-analysis looked at four randomised, double-blind, placebo-controlled trials of calcium supplementation (ranging from 1200-2000 mg/d) with the number of participants ranging from 194 to 1523 (total participants in calcium group=1487, total participants in placebo group=1497) (58). This study found an overall modest protective effect of calcium supplementation on recurrence of colorectal adenomas (random-effects risk ratio (RR) = 0.87, 95% CI: 0.77-0.98, I^2^=38.7%) (58). Another systematic review and meta-analysis included two trials comparing supplemental calcium (ranging from 1200-2000 mg/d) versus placebo for recurrence of adenomas, in 354-832 participants (total participants in calcium group=585, total participants in placebo group=601) (59). This analysis also found a risk reduction in developing an adenoma in the calcium supplementation arm compared to placebo (random-effects RR:0.82, 95%CI: 0.69–0.98, I^2^=0%) (59). Another systematic review of RCTs found contradictory results, with some studies finding increased risk or no effect, however, they were unable to perform a meta-analysis due to high heterogeneity between studies (60). A meta-analysis of cohort studies, including six studies with a total of 920,837 participants (including 8,839 colorectal cancer cases), also found a protective association of calcium supplementation with colorectal cancer risk (random effects RR per 300 mg/d = 0.91, 95% CI = 0.86-0.98, I^2^ = 67%) (61). A recent large prospective study of diet and colorectal cancer risk in 542,778 participants from the Million Women Study (12,251 incident cases over 16.6 years) found a strong inverse association for dietary calcium (RR per 300 mg/day = 0.83, 95% CI 0.77–0.89, p<1×10^-6^) (62). Serum calcium levels are tightly regulated and therefore may not be reflective of dietary intake (63, 64). This could explain why the magnitude of effect in this study, where we used instruments generated from serum levels, was much smaller than studies measuring the effect of dietary or supplemental calcium. As previous prevention trials have focussed on recurrent adenomas and, given the latency period from colorectal adenoma to carcinoma (i.e. up to 10 years) (65, 66), large, longer-term primary prevention trials are required to confirm the potential protective effects of calcium supplementation on colorectal cancer development.

There are some considerations to acknowledge when conducting MR studies of nutritional factors. Firstly, sample sizes of GWASs are often small and so there are limited genetic variants available to instrument traits. For example, when constructing an instrument for beta-carotene, only one SNP was available. Subsequent MR studies may also be underpowered to detect effects given the small effect sizes of genetic variants on micronutrients, often due to a small heritable component of dietary micronutrient levels. There is often a lack of biological understanding of how genetic instruments influence micronutrient levels, limiting our ability to understand if variants are likely to be valid instruments, and subsequently our interpretation of MR findings. Circulating levels of biomarkers may also not reflect cellular levels of these biomarkers and often do not correlate with dietary intake (67), therefore limiting the application of findings to dietary interventions.

We found little evidence for an effect of genetically-proxied inhibition of the protein targets of lipid-lowering medication (e.g. statins), on colorectal cancer risk, and effect estimates were not consistent in direction. This is in disagreement with previous conventional observational studies that have found that lipid-lowering medications are associated with reduced colorectal cancer risk (68, 69). This relationship is also supported by a recent MVMR study that found an adverse effect of LDL cholesterol on colorectal cancer, adjusted for high-density lipoprotein (HDL) cholesterol and triglycerides (70). However, in the current analysis, we may have been underpowered to detect an effect of lipid-lowering drug targets on colorectal cancer given the far fewer number of genetic variants available to instrument these traits as compared to LDL cholesterol levels.

One strength of this analysis was that when proxying the effect of drug targets, genetic variants were selected that were located within genes that encode the drug’s protein target, which should minimise risk of horizontal pleiotropy bias and reverse causation. For most of the protein drug targets, with the exception of ACE, we instrumented a downstream biomarker of that particular drug (e.g. for lipid-lowering drugs this was reduced LDL cholesterol levels, and for antihypertensives this was reduced systolic blood pressure (SBP)). An advantage of using these variants instead of genetic variants associated with circulating levels of proteins (i.e. protein quantitative trait loci (pQTL)) is that these biomarkers are further along the causal pathway to the outcome, so these variants are more likely to mimic the drug effect. For ACE, we used variants associated with circulating plasma levels of this protein (pQTLs), this approach is consistent with previously published work, investigating the effect of genetically proxied ACE inhibition on cancer risk (27). The top variant (rs4343) in the ACE instrument explains a significant proportion of the variance in levels of this protein (9%) and is also associated with systolic blood pressure.

There are lower rates of colorectal cancer in women as compared to men, and it has been suggested that sex hormones may play a role in this disparity (6, 71). However, we found little evidence for an effect of most of the hormones tested on colorectal cancer risk. We found evidence that progesterone has a protective effect on colorectal cancer risk in men, however the confidence intervals for this effect were wide, indicating uncertainty around the true estimate. This finding also does not explain the sex disparity observed in colorectal cancer rates. A RCT examining the effect of taking a daily combined oestrogen and progestin tablet on colorectal cancer risk in women initially found evidence for a protective effect on colorectal cancer risk (hazard ratio (HR): 0.63, 95% CIs: 0.43-0.92) (72). However, this study later found this finding was due to diagnostic delays and those in the treatment group were more likely to be diagnosed with advanced disease (73). This is in agreement with our findings, that showed little evidence for an effect of progesterone on colorectal cancer risk in women. This analysis was limited as we were unable to conduct further sensitivity analyses for these findings given the small number of SNPs available to instrument these hormones.

These analyses were restricted to individuals of European ancestry and therefore findings may not be generalisable to other ancestry groups. This is particularly relevant in the setting of therapeutic agents, given that they may have different pharmacokinetic/pharmacodynamic (PKPD) properties depending on germline pharmacogenomic variants that vary across populations (74, 75). In future, when GWAS data is available in different populations for the molecular traits examined here, there would be value in repeating these analyses using a more diverse cohort.

## CONCLUSION

These analyses found genetic evidence for a causal effect of several potential preventive agents on colorectal cancer risk. Reduced plasma ACE levels were robustly linked to increased colorectal cancer risk, and there was some evidence to support that elevated calcium and progesterone (in men) were inversely associated with colorectal cancer. However, given the limitations outlined here, further research is required to understand if our findings reflect the effect of supplementation or medications on colorectal cancer prevention, and whether they can be translated to clinical interventions. With new large-scale genetic and proteomic datasets becoming available, there is likely great potential for identifying novel targets for cancer prevention in future (76).

## Supporting information

STROBE-MR checklist

## ACKNOWLEDGMENTS

ASTERISK: We are very grateful to those without whom this project would not have existed. We also thank all those who agreed to participate in this study, including the patients and the healthy control persons, as well as all the physicians, technicians and students.

CCFR: The Colon CFR graciously thanks the generous contributions of their study participants, dedication of study staff, and the financial support from the U.S. National Cancer Institute, without which this important registry would not exist. The authors would like to thank the study participants and staff of the Seattle Colon Cancer Family Registry and the Hormones and Colon Cancer study (CORE Studies).

CLUE II: We thank the participants of Clue I and Clue II and appreciate the continued efforts of the staff at the Johns Hopkins George W. Comstock Center for Public Health Research and Prevention in the conduct of the Clue Cohort Studies.

Maryland Cancer Registry (MCR) Cancer data was provided by the Maryland Cancer Registry, Center for Cancer Prevention and Control, Maryland Department of Health, with funding from the State of Maryland and the Maryland Cigarette Restitution Fund. The collection and availability of cancer registry data is also supported by the Cooperative Agreement NU58DP007114, funded by the Centers for Disease Control and Prevention. Its contents are solely the responsibility of the authors and do not necessarily represent the official views of the Centers for Disease Control and Prevention or the Department of Health and Human Services.

COLON and NQplus: the authors would like to thank the COLON and NQplus investigators at Wageningen University & Research and the involved clinicians in the participating hospitals.

CORSA: We kindly thank all individuals who agreed to participate in the CORSA study. Furthermore, we thank all cooperating physicians and students and the Biobank Graz of the Medical University of Graz.

CPS-II: The authors express sincere appreciation to all Cancer Prevention Study-II participants, and to each member of the study and biospecimen management group. The authors would like to acknowledge the contribution to this study from central cancer registries supported through the Centers for Disease Control and Prevention’s National Program of Cancer Registries and cancer registries supported by the National Cancer Institute’s Surveillance Epidemiology and End Results Program. The authors assume full responsibility for all analyses and interpretation of results. The views expressed here are those of the authors and do not necessarily represent the American Cancer Society or the American Cancer Society – Cancer Action Network. Czech Republic CCS: We are thankful to all clinicians in major hospitals in the Czech Republic, without whom the study would not be practicable. We are also sincerely grateful to all patients participating in this study.

DACHS: We thank all participants and cooperating clinicians, and everyone who provided excellent technical assistance.

EDRN: We acknowledge all contributors to the development of the resource at University of Pittsburgh School of Medicine, Department of Gastroenterology, Department of Pathology, Hepatology and Nutrition and Biomedical Informatics.

EPIC: Where authors are identified as personnel of the International Agency for Research on Cancer/World Health Organization, the authors alone are responsible for the views expressed in this article and they do not necessarily represent the decisions, policy or views of the International Agency for Research on Cancer/World Health Organization.

EPICOLON: We are sincerely grateful to all patients participating in this study who were recruited as part of the EPICOLON project. We acknowledge the Spanish National DNA Bank, Biobank of Hospital Clínic–IDIBAPS and Biobanco Vasco for the availability of the samples. The work was carried out (in part) at the Esther Koplowitz Centre, Barcelona.

Harvard cohorts: The study protocol was approved by the institutional review boards of the Brigham and Women’s Hospital and Harvard T.H. Chan School of Public Health, and those of participating registries as required. We acknowledge Channing Division of Network Medicine, Department of Medicine, Brigham and Women’s Hospital as home of the NHS. The authors would like to acknowledge the contribution to this study from central cancer registries supported through the Centers for Disease Control and Prevention’s National Program of Cancer Registries (NPCR) and/or the National Cancer Institute’s Surveillance, Epidemiology, and End Results (SEER) Program. Central registries may also be supported by state agencies, universities, and cancer centers. Participating central cancer registries include the following: Alabama, Alaska, Arizona, Arkansas, California, Colorado, Connecticut, Delaware, Florida, Georgia, Hawaii, Idaho, Indiana, Iowa, Kentucky, Louisiana, Massachusetts, Maine, Maryland, Michigan, Mississippi, Montana, Nebraska, Nevada, New Hampshire, New Jersey, New Mexico, New York, North Carolina, North Dakota, Ohio, Oklahoma, Oregon, Pennsylvania, Puerto Rico, Rhode Island, Seattle SEER Registry, South Carolina, Tennessee, Texas, Utah, Virginia, West Virginia, Wyoming. The authors assume full responsibility for analyses and interpretation of these data.

Kentucky: We would like to acknowledge the staff at the Kentucky Cancer Registry. LCCS: We acknowledge the contributions of all who conducted this study which was originally reported as 10.1093/carcin/24.2.275.

NCCCS I & II: We would like to thank the study participants, and the NC Colorectal Cancer Study staff.

NSHDS investigators thank the Västerbotten Intervention Programme, the Northern Sweden MONICA study, the Biobank Research Unit at Umeå University and Biobanken Norr at Region Västerbotten for providing data and samples and acknowledge the contribution from Biobank Sweden, supported by the Swedish Research Council.

PLCO: The authors thank the PLCO Cancer Screening Trial screening center investigators and the staff from Information Management Services Inc and Westat Inc. Most importantly, we thank the study participants for their contributions that made this study possible.

Cancer incidence data have been provided by the District of Columbia Cancer Registry, Georgia Cancer Registry, Hawaii Cancer Registry, Minnesota Cancer Surveillance System, Missouri Cancer Registry, Nevada Central Cancer Registry, Pennsylvania Cancer Registry, Texas Cancer Registry, Virginia Cancer Registry, and Wisconsin Cancer Reporting System. All are supported in part by funds from the Center for Disease Control and Prevention, National Program for Central Registries, local states or by the National Cancer Institute, Surveillance, Epidemiology, and End Results program. The results reported here and the conclusions derived are the sole responsibility of the authors.

SEARCH: We thank the SEARCH team

SELECT: We thank the research and clinical staff at the sites that participated on SELECT study, without whom the trial would not have been successful. We are also grateful to the 35,533 dedicated men who participated in SELECT.

WHI: The authors thank the WHI investigators and staff for their dedication, and the study participants for making the program possible. A full listing of WHI investigators can be found at: https://s3-us-west-2.amazonaws.com/www-whi-org/wp-content/uploads/WHI-Investigator-Long-List.pdf

This manuscript is the result of funding in whole or in part by the National Institutes of Health (NIH). It is subject to the NIH Public Access Policy. Through acceptance of this federal funding, NIH has been given a right to make this manuscript publicly available in PubMed Central upon the Official Date of Publication, as defined by NIH.

## DATA AVAILABILITY

We obtained publicly available summary genetic association data for colorectal cancer from GECCO (https://www.ebi.ac.uk/gwas/publications/36539618). Approval was received to use restricted summary genetic association data from GECCO consortia after submitting a proposal to GECCO to access this data. We obtained publicly available summary genetic association data for all exposures, the GWAS PMID for each study used is presented in Table 1 and details for accessing summary data available can be found in each paper.

