## Supplementary material for "Investigating the causal effect of potential therapeutic agents for colorectal cancer prevention: a Mendelian randomization analysis": STROBE-MR checklist

**STROBE-MR checklist of recommended items to address in reports of Mendelian randomization studies**^1^ ^2^

| **Item No.** | **Section** | **Checklist item** | **Page No.** | **Relevant text from manuscript** |
| --- | --- | --- | --- | --- |
| 1 | **TITLE and ABSTRACT** | Indicate Mendelian randomization (MR) as the study’s design in the title and/or the abstract if that is a main purpose of the study | 1, 3 | “Title: Investigating the causal effect of previously reported therapeutic agents for colorectal cancer prevention: a Mendelian randomization analysis”  Abstract: “We used Mendelian randomization (MR), a genetic epidemiological method that can strengthen causal inference, to evaluate the effect of..” |
|  | **INTRODUCTION** |  |  |  |
| 2 | **Background** | Explain the scientific background and rationale for the reported study. What is the exposure? Is a potential causal relationship between exposure and outcome plausible? Justify why MR is a helpful method to address the study question | 4-5 | Paragraph 1-3 Introduction, discusses current issue relating to modifiable risk factors and screening programmes. Introduces therapeutic prevention and findings from conventional observational epidemiological studies. Introduces MR as the method used in this study to reassess these previously identified relationships. |
| 3 | **Objectives** | State specific objectives clearly, including pre-specified causal hypotheses (if any). State that MR is a method that, under specific assumptions, intends to estimate causal effects | 5 | “We used Mendelian randomization to reassess these previously identified observational relationships and provide evidence to support their causal nature” |
|  | **METHODS** |  |  |  |
| 4 | **Study design and data sources** | Present key elements of the study design early in the article. Consider including a table listing sources of data for all phases of the study. For each data source contributing to the analysis, describe the following: |  |  |
|  | a) | Setting: Describe the study design and the underlying population, if possible. Describe the setting, locations, and relevant dates, including periods of recruitment, exposure, follow-up, and data collection, when available. | 5-8 | We state we use a two-sample MR framework and that we restricted studies to those conducted in individuals of European ancestry |
|  | b) | Participants: Give the eligibility criteria, and the sources and methods of selection of participants. Report the sample size, and whether any power or sample size calculations were carried out prior to the main analysis | 5-8 | We used summary data from genome-wide association studies, so this information is reported elsewhere (all studies referenced). We report sample size of outcome study on page 6 and provide a table in the results section of exposure studies and sample sizes (page 9/10). We previously calculated power in our published analysis protocol for this study (https://doi.org/10.12688/wellcomeopenres.20861.2) |
|  | c) | Describe measurement, quality control and selection of genetic variants |  | Selection of genetic variants is described in our published protocol (<https://doi.org/10.12688/wellcomeopenres.20861.2>) measurement and quality control is described in publications for each GWAS used. |
|  | d) | For each exposure, outcome, and other relevant variables, describe methods of assessment and diagnostic criteria for diseases |  | Available in relevant paper of each GWAS |
|  | e) | Provide details of ethics committee approval and participant informed consent, if relevant | N/a | We used summary data only. Details of ethics and consent available in each paper |
| 5 | **Assumptions** | Explicitly state the three core IV assumptions for the main analysis (relevance, independence and exclusion restriction) as well assumptions for any additional or sensitivity analysis |  | The three core IV assumptions are described in detail in our published protocol as well as for a number of sensnitivy analysises (<https://doi.org/10.12688/wellcomeopenres.20861.2>) |
| 6 | **Statistical methods: main analysis** | Describe statistical methods and statistics used |  |  |
|  | a) | Describe how quantitative variables were handled in the analyses (i.e., scale, units, model) | 6 | We describe on page 6 how effect estimates were scaled, |
|  | b) | Describe how genetic variants were handled in the analyses and, if applicable, how their weights were selected | 7-8 | We describe using different LD thresholds to select genetic instruments for different classes of molecular traits |
|  | c) | Describe the MR estimator (e.g. two-stage least squares, Wald ratio) and related statistics. Detail the included covariates and, in case of two-sample MR, whether the same covariate set was used for adjustment in the two samples | 11 | we define odds ratio on page 11. Covariates are detailed in published protocol (<https://doi.org/10.12688/wellcomeopenres.20861.2>) |
|  | d) | Explain how missing data were addressed | n/a |  |
|  | e) | If applicable, indicate how multiple testing was addressed | 8 | We used an FDR correction 5% to account for multiple testing |
| 7 | **Assessment of assumptions** | Describe any methods or prior knowledge used to assess the assumptions or justify their validity | 9 | F-statistic and R-squared calculations are provided (relating to the “relevance” assumption) in Supplementary table 2, referenced on page 9 |
| 8 | **Sensitivity analyses and additional analyses** | Describe any sensitivity analyses or additional analyses performed (e.g. comparison of effect estimates from different approaches, independent replication, bias analytic techniques, validation of instruments, simulations) | 6-9 | Sensitivity analyses described on pages 6-9 and in published protocol (<https://doi.org/10.12688/wellcomeopenres.20861.2>) |
| 9 | **Software and pre-registration** |  |  |  |
|  | a) | Name statistical software and package(s), including version and settings used |  | Statistical software used is mentioned and referenced through this study and in the published protocol (<https://doi.org/10.12688/wellcomeopenres.20861.2>) |
|  | b) | State whether the study protocol and details were pre-registered (as well as when and where) | 5 | Protocol paper referenced in first paragraph of methods |
|  | **RESULTS** |  |  |  |
| 10 | **Descriptive data** |  |  |  |
|  | a) | Report the numbers of individuals at each stage of included studies and reasons for exclusion. Consider use of a flow diagram | 6-10 | Outcome study sample sizes reported throughout text and exposure study samples sizes reported in table on page 9/10 |
|  | b) | Report summary statistics for phenotypic exposure(s), outcome(s), and other relevant variables (e.g. means, SDs, proportions) | n/a | Not provided by individual genome-wide association studies |
|  | c) | If the data sources include meta-analyses of previous studies, provide the assessments of heterogeneity across these studies | 6 | “Between study heterogeneity was calculated using the *I^2^* statistic and variants with *I^2^* >65% were excluded.” |
|  | d) | For two-sample MR:  i.  Provide justification of the similarity of the genetic variant-exposure associations between the exposure and outcome samples  ii.  Provide information on the number of individuals who overlap between the exposure and outcome studies |  |  |
| 11 | **Main results** |  |  |  |
|  | a) | Report the associations between genetic variant and exposure, and between genetic variant and outcome, preferably on an interpretable scale | S | Reported in supplemtary tables 2 |
|  | b) | Report MR estimates of the relationship between exposure and outcome, and the measures of uncertainty from the MR analysis, on an interpretable scale, such as odds ratio or relative risk per SD difference | 11-14 | OR per SD increase/decrease, 95% CIs, for each exposure/outcome relationship presented in figures and throughout the text |
|  | c) | If relevant, consider translating estimates of relative risk into absolute risk for a meaningful time period | na | na |
|  | d) | Consider plots to visualize results (e.g. forest plot, scatterplot of associations between genetic variants and outcome versus between genetic variants and exposure) | 11-14 | All MR estimates presented in forest plots |
| 12 | **Assessment of assumptions** |  |  |  |
|  | a) | Report the assessment of the validity of the assumptions | 11-19 | F statistics provided in Supplementary table 2 (assessing “relevance assumption). Various sensitivity analyses conducted to assess “exclusion restriction” assumption |
|  | b) | Report any additional statistics (e.g., assessments of heterogeneity across genetic variants, such as *I^2^*, Q statistic or E-value) | na | na |
| 13 | **Sensitivity analyses and additional analyses** |  |  |  |
|  | a) | Report any sensitivity analyses to assess the robustness of the main results to violations of the assumptions | 11-19 | Sensitivity analyses and results reported throughout the text and in figures |
|  | b) | Report results from other sensitivity analyses or additional analyses | 11-19 | Sensitivity analyses and results reported throughout the text and in figures |
|  | c) | Report any assessment of direction of causal relationship (e.g., bidirectional MR) | 18 | Steiger filtering used to test causal direction of SNP effects |
|  | d) | When relevant, report and compare with estimates from non-MR analyses | 21-26 | This is done throughout the discussion |
|  | e) | Consider additional plots to visualize results (e.g., leave-one-out analyses) | 19 | Regional association plot to accompany colocalisation analyses |
|  | **DISCUSSION** |  |  |  |
| 14 | **Key results** | Summarize key results with reference to study objectives | 21-22 | First paragraph of discussion summarizes key results |
| 15 | **Limitations** | Discuss limitations of the study, taking into account the validity of the IV assumptions, other sources of potential bias, and imprecision. Discuss both direction and magnitude of any potential bias and any efforts to address them | 21-25 | Limitations discussed throughout discussion |
| 16 | **Interpretation** |  |  |  |
|  | a) | Meaning: Give a cautious overall interpretation of results in the context of their limitations and in comparison with other studies | 21-26 | Each finding is discussed individually in the context of limitations and other studies |
|  | b) | Mechanism: Discuss underlying biological mechanisms that could drive a potential causal relationship between the investigated exposure and the outcome, and whether the gene-environment equivalence assumption is reasonable. Use causal language carefully, clarifying that IV estimates may provide causal effects only under certain assumptions | 21-26 | Potential mechanisms are discussed throughout the discussion |
|  | c) | Clinical relevance: Discuss whether the results have clinical or public policy relevance, and to what extent they inform effect sizes of possible interventions | 26 | Clinical relevance discussed in conclusion |
| 17 | **Generalizability** | Discuss the generalizability of the study results (a) to other populations, (b) across other exposure periods/timings, and (c) across other levels of exposure | 21-26 | Issues discussed such as whether serum levels of biomarkers reflect dietary intake |
|  | **OTHER INFORMATION** |  |  |  |
| 18 | **Funding** | Describe sources of funding and the role of funders in the present study and, if applicable, sources of funding for the databases and original study or studies on which the present study is based | 26 | Funding described on page 26 |
| 19 | **Data and data sharing** | Provide the data used to perform all analyses or report where and how the data can be accessed, and reference these sources in the article. Provide the statistical code needed to reproduce the results in the article, or report whether the code is publicly accessible and if so, where | 27 | Data availability statement |
| 20 | **Conflicts of Interest** | All authors should declare all potential conflicts of interest | 27 | Competing interests statement |

This checklist is copyrighted by the Equator Network under the Creative Commons Attribution 3.0 Unported (CC BY 3.0) license.

1.	Skrivankova VW, Richmond RC, Woolf BAR, Yarmolinsky J, Davies NM, Swanson SA, et al. Strengthening the Reporting of Observational Studies in Epidemiology using Mendelian Randomization (STROBE-MR) Statement. JAMA. 2021;under review.

2.	Skrivankova VW, Richmond RC, Woolf BAR, Davies NM, Swanson SA, VanderWeele TJ, et al. Strengthening the Reporting of Observational Studies in Epidemiology using Mendelian Randomisation (STROBE-MR): Explanation and Elaboration. BMJ. 2021;375:n2233.
